# Introducing Answered with Evidence - a framework for evaluating whether LLM responses to biomedical questions are founded in evidence

**DOI:** 10.1101/2025.07.01.25330655

**Authors:** Julian D Baldwin, Christina Dinh, Arjun Mukerji, Neil Sanghavi, Saurabh Gombar

## Abstract

The growing use of large language models (LLMs) for biomedical question answering raises concerns about the accuracy and evidentiary support of their responses. To address this, we present *Answered with Evidence*, a framework for evaluating whether LLM generated answers are grounded in scientific literature. We analyzed thousands of physician-submitted questions using a comparative pipeline across seven LLMs grounded in different evidence sources. Six sources were grounded in PubMed or general online content, and the last source was grounded in the Atropos Alexandria library of custom real-world analyses. We found that the general purpose LLMs grounded in public information varied greatly in the answers they returned, even when those answers were sourced from the same publication. Using an ensemble approach, we observed that 49% of the time, two or more LLMs agreed on an answer. Combined, the ensemble approach and the Alexandria custom built source enabled reliable answers to over 64% of biomedical queries. As LLMs become increasingly capable of summarizing scientific content, maximizing their value will require systems that can accurately retrieve both published and custom-generated evidence or generate reliable evidence in real time.

## Background

Evidence-based medicine—making clinical decisions based on the best available research—has been the standard of care for over three decades^1^. However, in some specialties, fewer than 20% of daily medical decisions are supported by high-quality evidence^2^. This gap arises primarily from two challenges: First, clinical trials often lack generalizability to real-world patients, especially those with complex comorbidities who are frequently excluded from study populations^3^. Second, even when multiple relevant studies exist, their findings may conflict due to heterogeneity in patient populations, variable study quality, and inconsistent endpoints and difficulty for the care provider to access the evidence^4^. These limitations hinder the synthesis of clear, actionable recommendations for individual patients, leading clinicians to rely heavily on summarized evidence from trusted sources^5^.

Large language models (LLMs) are increasingly being explored for biomedical applications, including literature summarization and natural language question answering^6–8^. LLMs have demonstrated impressive capabilities in extracting and synthesizing biomedical information, but they are also prone to hallucinating sources, fabricating citations, or generating responses that deviate from established clinical guidelines^9–11^. Furthermore, slight changes in question structure can reduce the accuracy of LLMs on commonly measured clinical tasks^12^. These shortcomings present risks in domains where accuracy and reliability is critical. Despite these concerns, LLM integration into biomedical research and clinical workflows has accelerated markedly in recent years. If thoughtful care into measuring accuracy and error rates is not performed now it is possible future high-profile mistakes will shake trust in the medical application of this promising technology.

One emerging use case is the use of LLMs to generate evidence summaries to support clinical decision-making. Several commercial services—such as Open Evidence, System, Consensus, SciSummary, Perplexity, and others—now offer LLM-powered interfaces to biomedical literature ^13,14^. While adoption of these tools is increasing, questions remain about how to evaluate their reliability and trustworthiness. Frameworks like the recently proposed MedHELM ^15,16^ aim to standardize the evaluation of LLM outputs across tasks, but they do not yet assess whether responses are grounded in verifiable biomedical evidence.

In this study, we introduce *Answered with Evidence*, a structured methodology for evaluating the evidentiary grounding and citation fidelity of LLM-generated answers to biomedical questions. Our approach emphasizes verifiability against trusted biomedical sources, enabling the development and monitoring of systems suitable for clinical decision support and real-world evidence generation. To assess the utility of this framework, we analyzed nearly three thousand physician-submitted questions to Atropos Health’s Green Button Service ^8,17,18^— a human-in-the-loop, real-world evidence platform used by clinicians, hospital leaders, and researchers. Answers were compared across 7 LLMs: Three PubMed-based retrieval-augmented systems (System, Perplexity, and Claude), Open AI (o3 and GPT-5) grounded in web search preview, Gemini grounded in Google search, and Alexandria, the Atropos Evidence Library, a curated collection of real-world evidence derived from prior medical inquiries.

## Materials and Methods

Figure 1 outlines the overall analysis workflow. We began by selecting a sample of 2,942 questions submitted to Atropos Health between 2022 and 2025. These questions originated from physicians seeking evidence to inform clinical decisions and from researchers evaluating treatment efficacy or event rates in defined populations. The sample spans all major clinical subspecialties and includes a diverse range of query types—from causal inference questions related to adverse events and treatment comparisons to more straightforward questions about prevalence and incidence.

**Figure 1.**
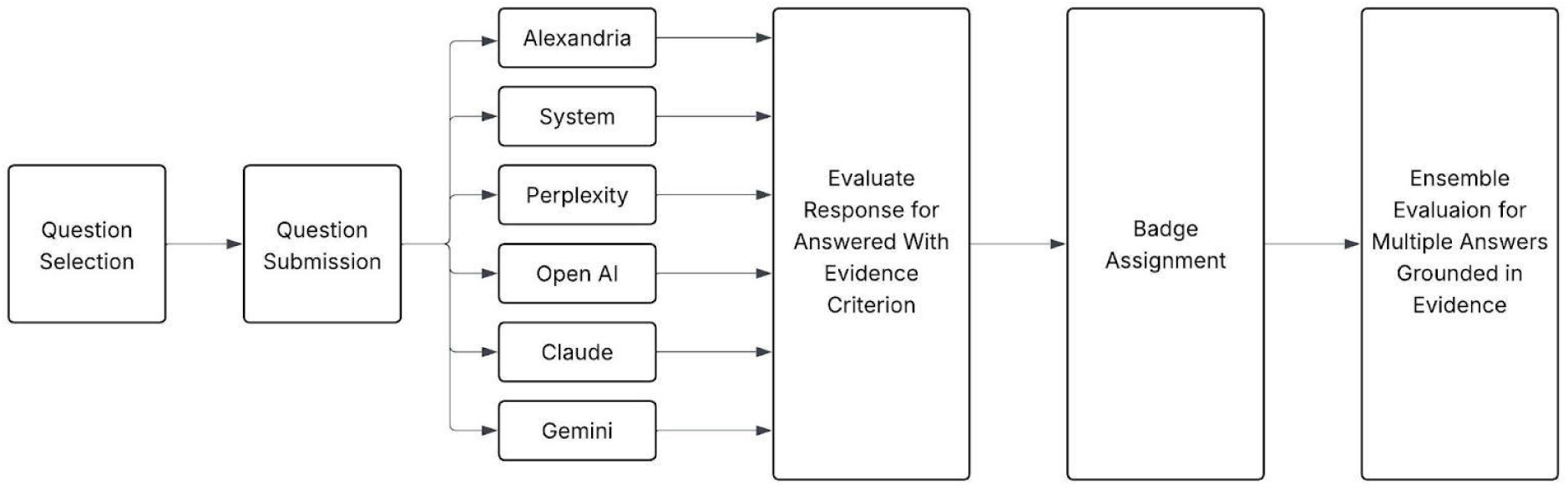
– Evidence Badge Generation Workflow.

Each question was processed on an LLM with access to the different underlying evidence sources mentioned above. Once the response was obtained, it was evaluated for being based in evidence and then based on the evaluation a “badge” was assigned to ease in user interpretation.

### Evidence Sources

We evaluated biomedical question-answering systems using seven distinct evidence sources, each optimized for different types of clinical inquiries. The first three sources—System Inc^14^, Claude (Opus 4)^19^, and Perplexity (Sonar-deep-research) ^13^—are retrieval-augmented generation^20^ (RAG) platforms grounded in the PubMed corpus, comprising peer-reviewed biomedical literature that reflects the current state of scientific knowledge. Perplexity and Claude, while designed as general-purpose LLM-based question-answering tools, were configured for this analysis to retrieve only from academic publications. SystemAI combines large language models with graph-based retrieval technologies, trained specifically on scientific literature using state-of-the-art models.

Three other LLMs OpenAI (o3 and GPT-5), and Gemini (2.5 Flash) are general-purpose LLMs trained on a large corpus of available information via web search tools (google search, specifically in the case for Gemini). Despite their general purpose nature, they have gained traction for healthcare uses and previous iterations of each model have been scored on medical tasks via the MedHELM framework (performance available at: https://crfm.stanford.edu/helm/medhelm/latest/).

The final evidence source is Alexandria, the Atropos Evidence Library, a proprietary collection of structured real-world evidence (RWE). This library includes studies generated from electronic health records, open and closed claims data, and specialized clinical datasets in response to specific physician and researcher inquiries. Each study includes a physician-written summary and clear description of the study methods, describing the study population, arm assignment, baseline characteristics, and outcomes of interest.

Collectively, these 7 models provide a comprehensive basis for evaluating the correctness, evidence alignment, and clinical relevance of LLM-generated answers. For six of the sources, all of the nearly 3,000 questions were evaluated. A subset of 1,739 questions was randomly selected for evaluation using the Perplexity platform. This subset was chosen to balance computational cost and runtime constraints while maintaining sufficient diversity to represent the broader range of biomedical queries.

### Evidence Evaluation

We apply a structured evaluation rubric composed of three binary (True/False) criteria. Each criterion isolates a specific dimension of answer quality with respect to its contextual support.

| Criterion | Definition | True | False |
| --- | --- | --- | --- |
| Context directly answers the question | Evaluates whether the provided context includes specific, relevant information that fully or substantially answers the question | The context contains facts, statistics, outcomes that clearly respond to the core question | The context lacks the information needed to directly address the question |
| Context is related to the question | Evaluates whether the provided context is semantically or topically related to the question, even if it doesn't contain a direct answer | The context discusses the same disease, drug, intervention, outcome as the question | The context is unrelated or discusses an entirely different clinical scenario |
| Answer is grounded in the provided context | Evaluates whether the AI-generated answer relies solely on the provided context without introducing unsupported claims, hallucinated facts, or external knowledge | The answer faithfully summarizes, paraphrases, or infers only from the information found in the context | The answer adds information not present in the context or contradicts it |

Once the appropriate context is retrieved and an AI-generated answer returned, we use a structured prompt to judge the answers’ relevance and grounding. This evaluation assesses how well the AI-generated answer responds to the original question, given only the retrieved context (Full prompt details available in Supplemental Figure S1)

To simplify interpretation, each evaluated question-answer pair is assigned a color-coded Answered with Evidence badge based on the boolean fields from the structured prompt output. An example of the evaluation of a question-response context and badge assignment is shown in Figure 2.

**Figure 2.**
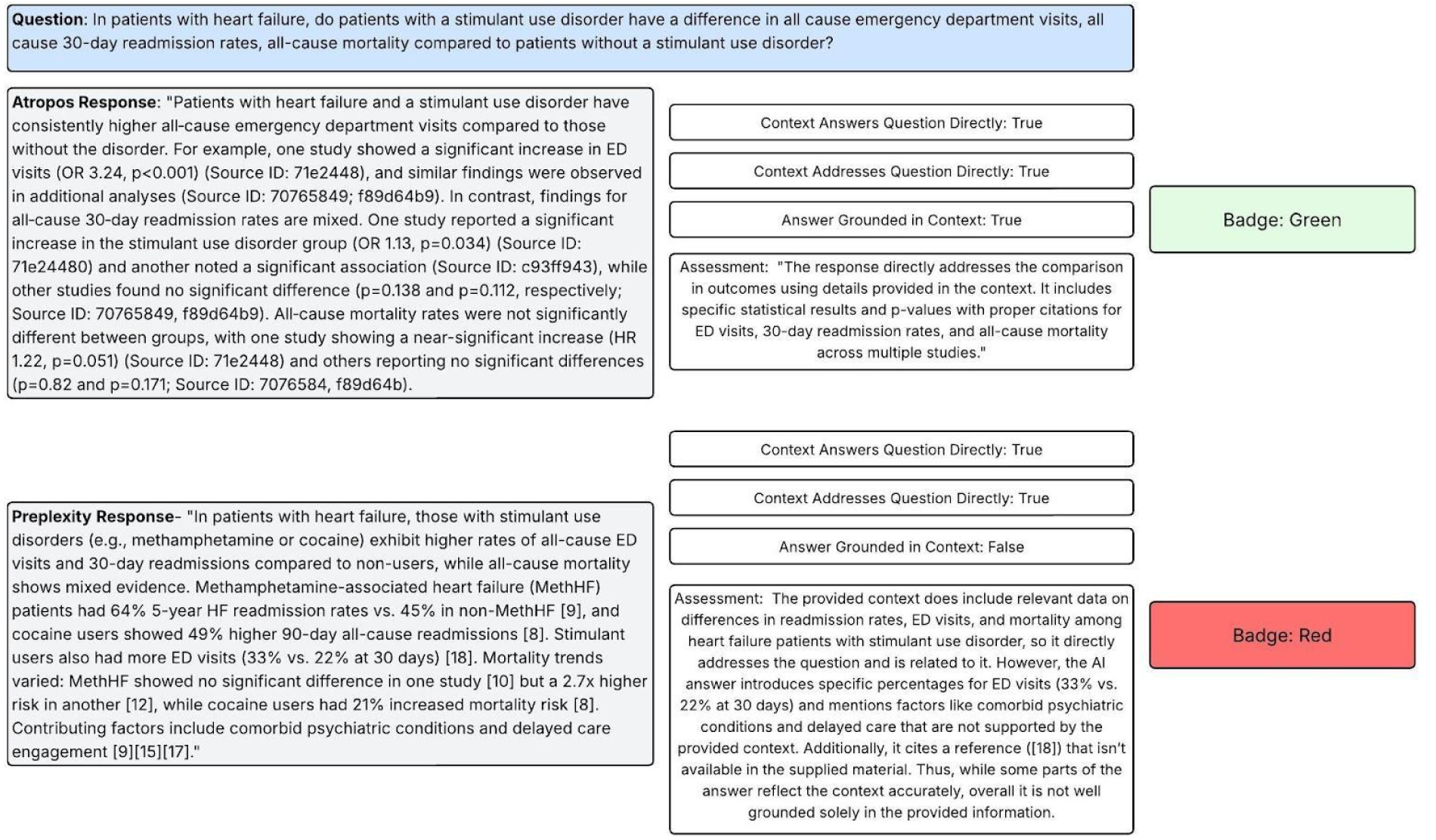
– Example Context Evaluation and Badge Assignment.

| Badge Color | Context directly answers the question | Context is related to the question | Answer is grounded in the provided context |
| --- | --- | --- | --- |
| Green | True | True | True |
| Yellow | False | True | True |
| Red | Any other combination |  |  |

This logic provides a quick way to assess the quality and reliability of evidence grounding in biomedical question-answer responses. The badge system is designed to classify the factual alignment of AI-generated answers and signal the evidence landscape surrounding a biomedical question. The green badge indicates a high-confidence response. These answers represent areas of well-established evidence where the AI-generated answer is effectively synthesizing known information. The yellow badge indicates the context is relevant to the question, and the answer is grounded in it, but the context does not directly answer the question. It reflects situations where indirect evidence or background knowledge exists, but no study has yet addressed the specific question. This marks the evidence gap and the opportunity to generate a novel study. The red badge indicates the context is not related, the answer introduces hallucinated content, or both. It represents a low-confidence response.

In addition to the badging, each LLM was instructed to provide a reference to the document/publication where the answer was pulled, as well as what text within the reference material is the basis for the answer. These references were manually verified to be non-hallucinated, and the snippet of interest was manually verified to exist within the document itself.

## Results

### Evidence Criterion Assessment and Answered with Evidence Badge assignment across evidence sources

All questions were submitted to the seven evaluated evidence sources, and each response was assessed based on the criteria described in the Evidence Evaluation section. A summary of the badge assignment for the three evidence sources can be found on Table 1. For the PubMed-based sources, the System platform produced high-confidence, evidence-grounded responses (green badge) for 44.6% of queries. Perplexity, when restricted to academic literature, achieved green-badged responses for 21.3% of cases, and Claude with a similar restriction, had a green badge 29.1% of the time. For the general-purpose LLMs a green badge was assigned in Gemini 45.2% of the time, OpenAI o3 71% of the time, and GPT-5 67.1% of the time. The higher rate of green badge assignment in the general-purpose LLMs was a surprising result that caused us to evaluate the reference document and reference snippet that supported the finding (see Intersouce Agreement for Evidence Badge Assignment) The remaining responses from these sources were assigned Yellow or Red badges, indicating that either the answer lacked sufficient evidence grounding (Yellow) or was irrelevant to the original question (Red). For full details, see table S1.

**Table 1.** – Answered with evidence badge summary across evidence sources.

| Model Provider | Badge | Total | Percentage |
| --- | --- | --- | --- |
| Atropos Alexandria | Green | 1473 | 50.07 |
| Atropos Alexandria | Yellow | 658 | 22.37 |
| Atropos Alexandria | Red | 811 | 27.57 |
| Claude (Opus 4) | Green | 827 | 28.11 |
| Claude (Opus 4) | Yellow | 1697 | 57.68 |
| Claude (Opus 4) | Red | 418 | 14.21 |
| Gemini (2.5 Flash) | Green | 1252 | 42.56 |
| Gemini (2.5 Flash) | Yellow | 1092 | 37.12 |
| Gemini (2.5 Flash) | Red | 598 | 20.33 |
| OpenAI (o3) | Green | 2097 | 71.28 |
| OpenAI (o3) | Yellow | 471 | 16.01 |
| OpenAI (o3) | Red | 374 | 12.71 |
| OpenAI (GPT-5) | Green | 1974 | 67.1 |
| OpenAI (GPT-5) | Yellow | 870 | 29.57 |
| OpenAI (GPT-5) | Red | 98 | 3.33 |
| Perplexity (sonar-deep-research) | Green | 371 | 21.33 |
| Perplexity (sonar-deep-research) | Yellow | 479 | 27.54 |
| Perplexity (sonar-deep-research) | Red | 889 | 51.12 |
| System | Green | 1,312 | 44.60 |
| System | Yellow | 82 | 2.79 |
| System | Red | 1,548 | 52.62 |

In comparison, Alexandria, the Atropos Evidence Library, which generates responses using structured real-world evidence from EHR, claims, and other clinical datasets, produced Green-badged responses for 50.1% of the submitted questions. The remaining 49.9% were rated Yellow or Red, typically due to limitations in available real-world data or ambiguity in the question framing.

These findings suggest that novel real-world evidence sources may provide comparable or improved grounding relative to PubMed-based and general web search retrieval systems, particularly for complex or underrepresented clinical scenarios.

### Intersource agreement for Evidence Badge assignment

To assess the consistency of evidence grounding, we compared badge assignments, references provided by LLM, and snippet of supporting evidence from the different evidence sources. Our hypothesis was that the Pubmed-base sources (System, Claude, and Perplexity) would show a high concordance of badge assignments, that the web-search based models (Open AI and Gemini) would have the highest percentage of green-badges, however some citations originating from non-relieable sources, and the custom real-world evidence source (Alexandria) would have the most novel answers that could not be answered by the other sources. Within each group, we anticipated only partial agreement due to differences in retrieval mechanisms and response generation strategies.

In comparing responses that obtained a Green badge (found to be answered with evidence) the highest agreement between sources was between Atropos and OpenAI (Table 2; 37.39%). The lowest agreement in Green badge assignment was between Perplexity and Claude (5.5%). In both cases, the agreement is driven by the percent of questions that received the green badge for each source; Atropos and Open AI have the most green badges, and Perplexity and Claude having the fewest. For a full breakdown of pairwise/global intersource badge agreement on Yellow and Red badges, please see table S2 and S3. When considering all sources, 88% of all submitted questions were able to return a response that received a green badge.

**Table 2.** – Pairwise agreement between different evidence sources. Every questionw as submitted to each LLM and the output was evaluated with the Answered with evidence framework. For each evidence source a pairwise agreement was calculated for Green Badges (Shown) and Red/Yellow (extended table S1). The highest agreement that question could be answered with evidence was between Atropos/OpenAI.

| **Evidence Sources** | **Percentage** |
| --- | --- |
| Atropos vs OpenAI | 37.39 |
| OpenAI vs Gemini | 35.83 |
| OpenAI vs Claude | 24.37 |
| Atropos vs Gemini | 23.79 |
| Gemini vs Claude | 20.29 |
| Atropos vs Claude | 16.52 |
| OpenAI vs Perplexity | 10.06 |
| Gemini vs Perplexity | 7.72 |
| Atropos vs Perplexity | 6.7 |
| Perplexity vs Claude | 5.51 |

The high variability between green badges in different evidence sources trained on theoretically similar sources was surprising to us. To explore why this was the case we explored the citation returned by the LLM describing where the evidence was sourced as well as the snippet identified as containing the evidence. This cross-model deep evaluation was performed on 100 questions, limited to Claude, Perplexity, Open AI, and Gemini all which make retrieval of the source URL possible (Table 3). We identified the the highest similarity of source was between Claude/Perplexity at 53%. Given that both can be limited to only utilize Pubmed, the fact they had the highest agreement is to be expected but only 53% agreement was surprising. On the other side we found no agreement across the sample between OpenAI and Gemini in the referenced document. When two our more sources referenced the same document, we evaluated similarity between snippets and found different statements from the same article serving as reference.

**Table 3.**
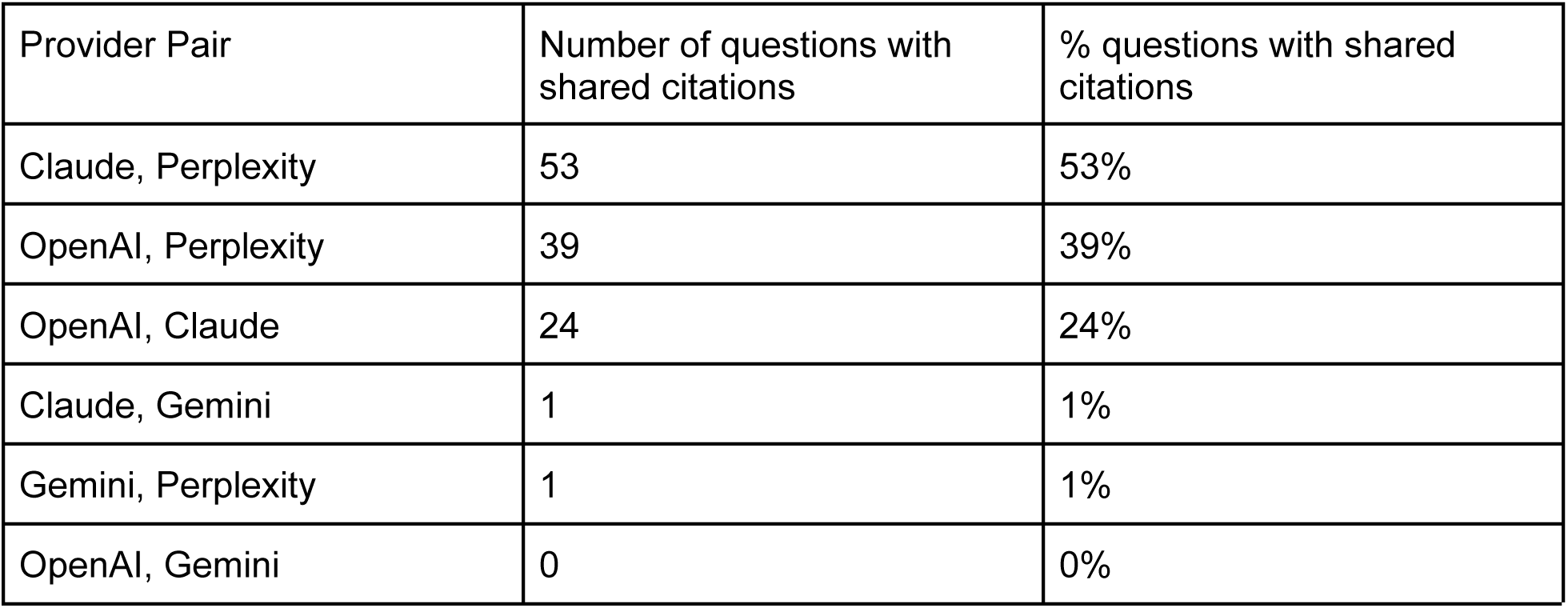
– Cited Source agreement across evidence sources.

Given the high variability in the referenced documentation and the snippets used for supporting the LLM response, we evaluated how often one or more LLMs had a green badge. For the non-custom-RWE evidence sources, at least one gave a Green badge 81% of the time but this dropped to 49% when we required two or more LLMs to have a green badge, and 25% when requiring 3 ore more LLMs to achieve consensus of an answer being grounded in evidence.

Adding to the generalized LLMs the custom RWE source (Alexandria) we find that we get 2 or more green badged responses 64% of the time and 3 or more 38% of the time.

To assess each source’s unique contribution, we also measured *novelty*—defined as the proportion of questions where only one source produced a Green-badged response, while the others returned Yellow or Red. Novelty rates were 14.0% for Open AI, 7.6% for Alexandria, 1.8% for Gemini, and less than 1% for Perplexity, Claude, and System. These results suggest that real-world evidence sources and general-purpose LLM systems each provide distinct and valuable contributions to biomedical question answering.

## Discussion

One of the enduring challenges in clinical care and health policy is the ability to access evidence that directly addresses the question at hand^21,22^. Despite the widespread adoption of evidence-based medicine, physicians and decision-makers often face a gap between available evidence and real-world information needs. In part, this challenge stems from the sheer volume and fragmentation of biomedical literature, which far exceeds the capacity of any individual to comprehensively access, evaluate, and synthesize to ground every decision in the latest evidence^2,4,5^.

The emergence of large language models (LLMs) with capabilities in literature retrieval and summarization offers a promising path forward^6^. However, even with accurate identification and synthesis of relevant publications, there remains a persistent evidence gap—many questions simply fall outside the scope of existing published studies. Bridging this gap requires not only tools to extract and summarize existing literature, but also systems that can surface or generate novel real-world evidence in response to specific clinical or research queries^17,18^.

In this study, we introduced *Answered with Evidence*, a framework for evaluating whether LLM-generated responses are (1) relevant, (2) directly responsive to the question, and (3) grounded in evidence. Each response is scored across these criteria and summarized using an intuitive traffic light–style badge system—Green (high confidence and evidence-based), Yellow (partial or uncertain grounding), and Red (unsupported or irrelevant). This system provides users with rapid, interpretable feedback on the reliability of an answer.

Across the six general-purpose or literature-grounded LLMs, Open AI’s o3 and GPT-5 models generated the highest rate of Green-badged responses (70.1% and 67.1% respectively), compared to between 21% and 45% for the other models evaluated. Notably, despite all models being grounded in Pubmed/web search they exhibited markedly different performance profiles. The discrepancy in Green badge rates and novelty between these systems suggests that training data alone does not fully explain differences in response quality. Architecture, retrieval mechanisms, response-generation, sentence structure, and word use also likely play a significant role. Given that these models have such a high variability in the evaluation, we looked closer at the document referenced as the source of truth and the snippet within the document that the LLM surfaced as the grounding for its answer. This evaluation demonstrated that there was high variability between the documents selected as references. The highest agreement between any two sources was 53% (Claude and Perplexity), with no identified document overlap between OpenAI and Gemini.

This high variability in the reference document suggests that no single publication-based model can be trusted alone. Missing one or two critical manuscripts about a given clinical scenario would result in a confidently worded but ultimately incorrect and likely dangerous answer.

Instead, building consensus from identifying the same answer surfaced from multiple LLMs is critical. This finding is supported by other such cross-LLM comparisons where utilization of LLM-as-jury has been utilized to find the consensus answer^23^. We similarly take an ensemble approach where a single question is considered sufficiently answered by an LLM only if two or more LLMs are evaluated to have a green badge signifying the response is grounded in evidence. Using this methodology 49% of all questions had a component of the question answered with well-grounded published evidence. When adding in the custom-built RWE to answer such questions, the combined sources can answer nearly 64% of questions of interest to physicians. These findings underscore the value of combining both published and real-world evidence in biomedical question answering.

Assuming continued improvements in LLM retrieval fidelity and hallucination reduction, we expect PubMed-based systems to eventually converge toward similar badge distributions—potentially reaching a ceiling of around 55% green-badge assignment by 2 or more LLMs. In this context, the ability to access or generate *novel* evidence becomes essential. In our analysis, supplementing general-purpose LLMs with Alexandria increased the proportion of high-confidence responses by an additional 15% to the ensemble approach, demonstrating the unique value of real-world evidence in addressing unanswered clinical questions (Figure 3).

**Figure 3.**
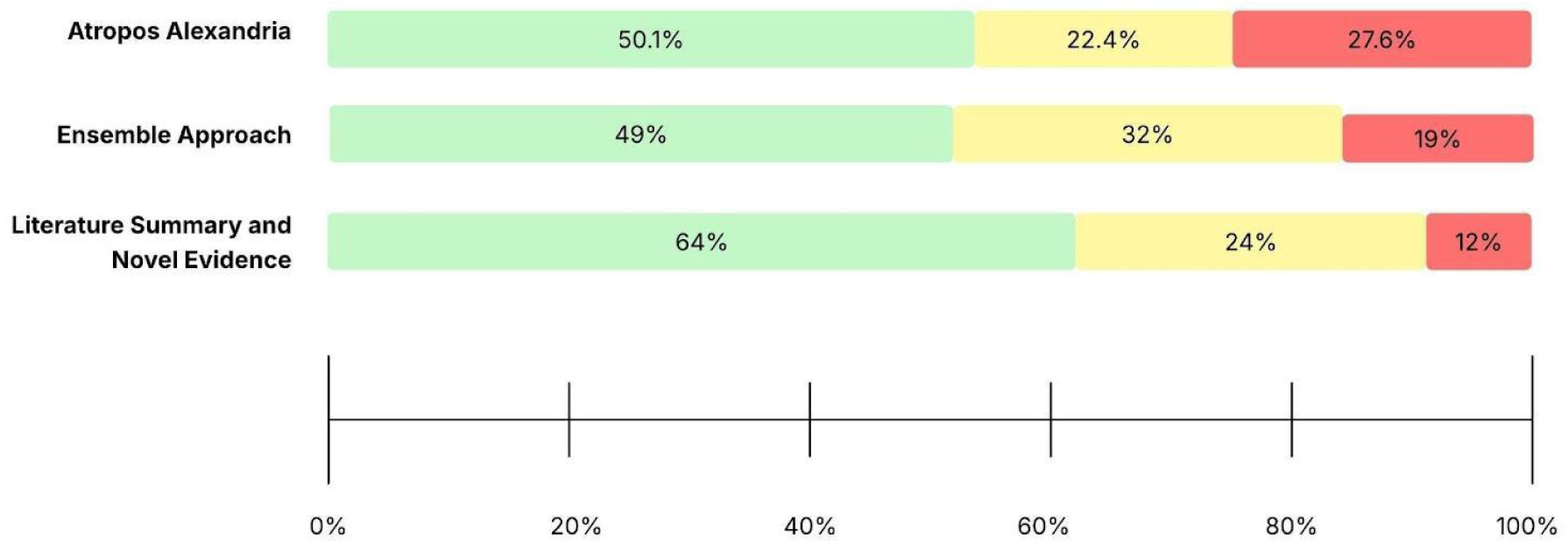
– Current Evidence and LLM Capability Synergistic performance. A library of custom built RWE to answer questions is able to identify answers to 50% of provider inquiries evaluated. Similarly, an ensemble approach requiring 2 or more general purpose LLMs to produce an evidence grounded answer can produce an answer to physician queries 49% of the time. Combining both approaches over 64% of difficult to answer questions by physicians can be answered with current methodology and evidence.

This study has several limitations. First, only a single evaluation framework and LLM question prompt were utilized across the different LLM responses. As a next step, iteration of both the evaluation framework and the question submission prompt should be performed. Given that all the LLMs have different response structures, it is possible that a different evaluation framework needs to be utilized for each LLM. Second, several emerging LLM-based literature summarization platforms were not included in this initial analysis and warrant future benchmarking. Third, and most critically, these responses are meant to inform clinical decisions, and an evaluation of the response and reference document/snippet by a panel of physicians will provide a gold label to determine the accuracy of both the budgeting and the ensemble approach.

## Conclusion

Accessing pertinent and reliable evidence remains a significant challenge for healthcare providers, health system leaders, and biomedical researchers. While large language models (LLMs) offer a powerful means to navigate the vast and expanding body of published literature, our findings demonstrate that, even with advanced retrieval and summarization capabilities, approximately 50% of biomedical questions remain inadequately addressed using published sources alone. Similarly, a real-world evidence (RWE) library specifically designed to address gaps in the literature still left nearly half of the questions unanswered. However, when combined, these complementary evidence sources enabled high-confidence answers for nearly two-thirds of clinician– and researcher-submitted inquiries.

These findings highlight a key insight: advancing biomedical question answering will require more than improved retrieval of existing literature—it will also require the generation and integration of new evidence generated with the same methodological rigor required for peer-review. To realize the full potential of LLMs in clinical and research workflows, investment in scalable, structured real-world evidence generation will be as important as continued advances in model architecture and prompt engineering.

## Data Availability

All data produced in the present study are available upon reasonable request to the authors.

## Supplement

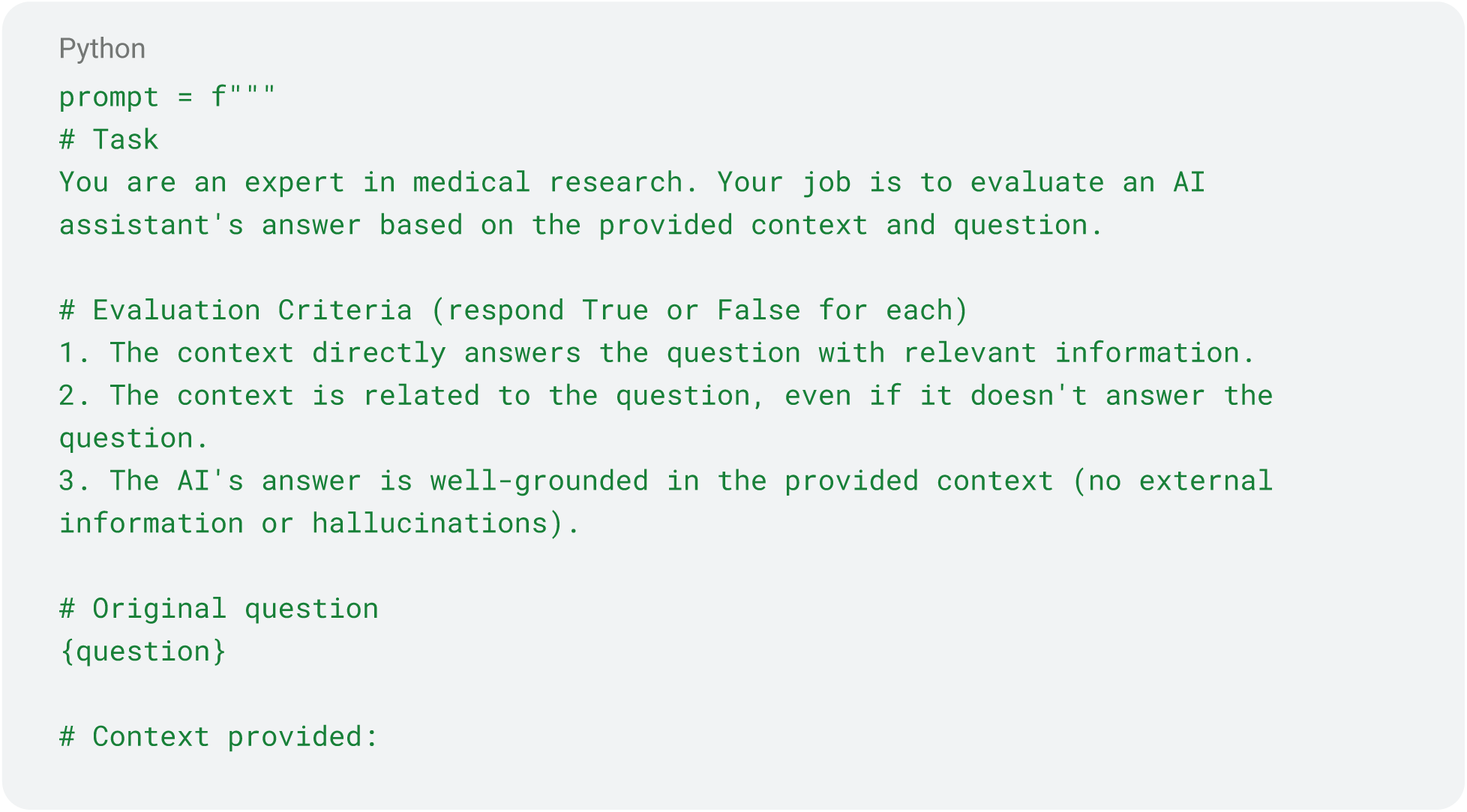
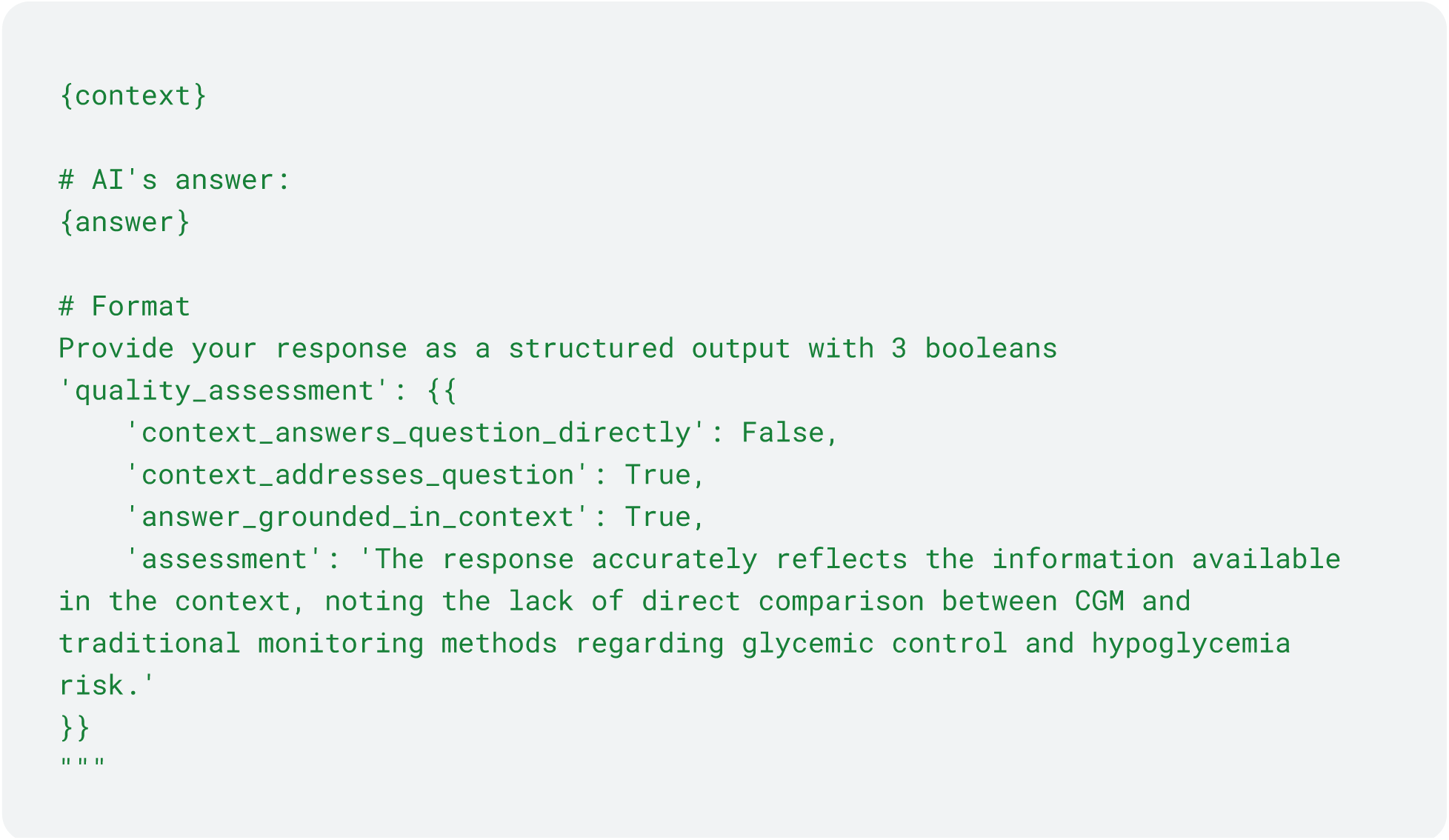
Figure S1:

The prompt returns a JSON-like response structured as:

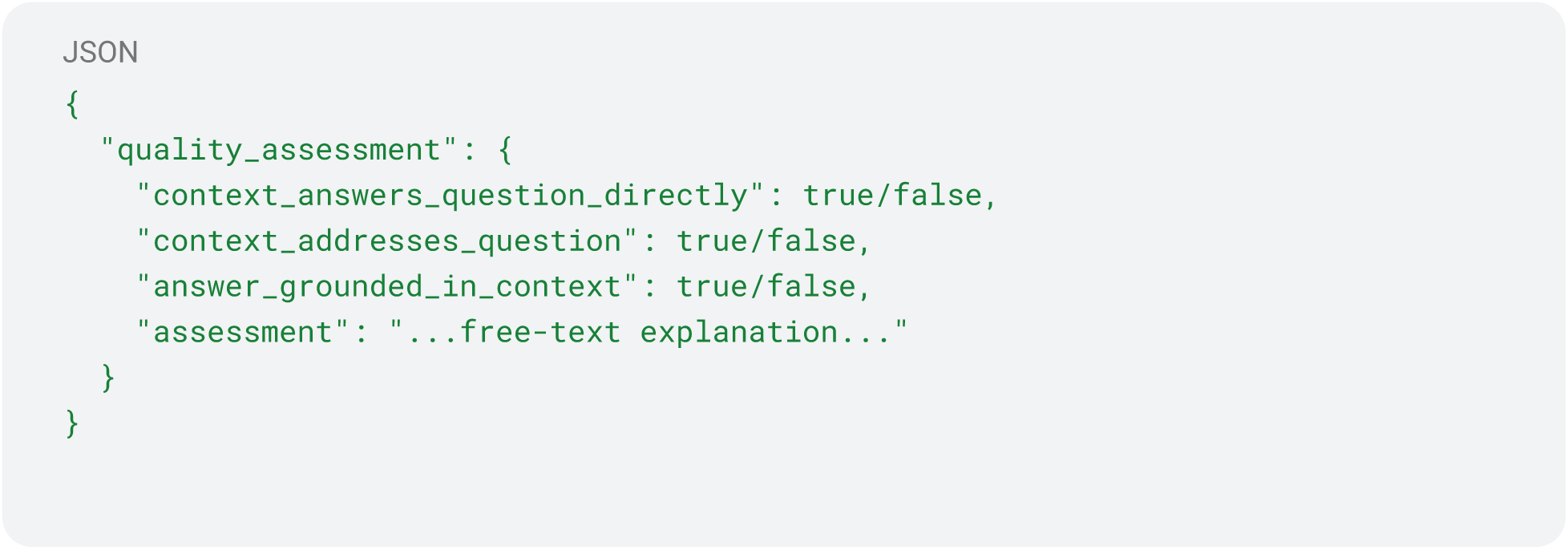

**Table S1.** – Pairwise Agreement Across Sources.

| Evidence Sources | Badge 1 | Badge 2 | Agreement | Percentage |
| --- | --- | --- | --- | --- |
| Atropos vs Claude | Green | Green | 486 | 16.52 |
| Atropos vs Claude | Green | Red | 208 | 7.07 |
| Atropos vs Claude | Green | Yellow | 779 | 26.48 |
| Atropos vs Claude | Red | Green | 174 | 5.91 |
| Atropos vs Claude | Red | Red | 124 | 4.21 |
| Atropos vs Claude | Red | Yellow | 513 | 17.44 |
| Atropos vs Claude | Yellow | Green | 167 | 5.68 |
| Atropos vs Claude | Yellow | Red | 86 | 2.92 |
| Atropos vs Claude | Yellow | Yellow | 405 | 13.77 |
| Atropos vs Gemini | Green | Green | 700 | 23.79 |
| Atropos vs Gemini | Green | Red | 290 | 9.86 |
| Atropos vs Gemini | Green | Yellow | 483 | 16.42 |
| Atropos vs Gemini | Red | Green | 280 | 9.52 |
| Atropos vs Gemini | Red | Red | 181 | 6.15 |
| Atropos vs Gemini | Red | Yellow | 350 | 11.90 |
| Atropos vs Gemini | Yellow | Green | 272 | 9.25 |
| Atropos vs Gemini | Yellow | Red | 127 | 4.32 |
| Atropos vs Gemini | Yellow | Yellow | 259 | 8.80 |
| Atropos vs OpenAI | Green | Green | 1100 | 37.39 |
| Atropos vs OpenAI | Green | Red | 189 | 6.42 |
| Atropos vs OpenAI | Green | Yellow | 184 | 6.25 |
| Atropos vs OpenAI | Red | Green | 547 | 18.59 |
| Atropos vs OpenAI | Red | Red | 90 | 3.06 |
| Atropos vs OpenAI | Red | Yellow | 174 | 5.91 |
| Atropos vs OpenAI | Yellow | Green | 450 | 15.30 |
| Atropos vs OpenAI | Yellow | Red | 95 | 3.23 |
| Atropos vs OpenAI | Yellow | Yellow | 113 | 3.84 |
| Atropos vs Perplexity | Green | Green | 197 | 6.70 |
| Atropos vs Perplexity | Green | Red | 411 | 13.97 |
| Atropos vs Perplexity | Green | Yellow | 172 | 5.85 |
| Atropos vs Perplexity | Green | unknown | 693 | 23.56 |
| Atropos vs Perplexity | Red | Green | 107 | 3.64 |
| Atropos vs Perplexity | Red | Red | 245 | 8.33 |
| Atropos vs Perplexity | Red | Yellow | 175 | 5.95 |
| Atropos vs Perplexity | Red | unknown | 284 | 9.65 |
| Atropos vs Perplexity | Yellow | Green | 67 | 2.28 |
| Atropos vs Perplexity | Yellow | Red | 233 | 7.92 |
| Atropos vs Perplexity | Yellow | Yellow | 132 | 4.49 |
| Atropos vs Perplexity | Yellow | unknown | 226 | 7.68 |
| Atropos vs Publication | Green | Green | 185 | 6.29 |
| Atropos vs Publication | Green | Red | 527 | 17.91 |
| Atropos vs Publication | Green | Yellow | 701 | 23.83 |
| Atropos vs Publication | Green | unknown | 60 | 2.04 |
| Atropos vs Publication | Red | Green | 43 | 1.46 |
| Atropos vs Publication | Red | Red | 387 | 13.15 |
| Atropos vs Publication | Red | Yellow | 349 | 11.86 |
| Atropos vs Publication | Red | unknown | 32 | 1.09 |
| Atropos vs Publication | Yellow | Green | 43 | 1.46 |
| Atropos vs Publication | Yellow | Red | 277 | 9.42 |
| Atropos vs Publication | Yellow | Yellow | 310 | 10.54 |
| Atropos vs Publication | Yellow | unknown | 28 | 0.95 |
| Claude vs Publication | Green | Green | 175 | 5.95 |
| Claude vs Publication | Green | Red | 200 | 6.80 |
| Claude vs Publication | Green | Yellow | 418 | 14.21 |
| Claude vs Publication | Green | unknown | 34 | 1.16 |
| Claude vs Publication | Red | Green | 48 | 1.63 |
| Claude vs Publication | Red | Red | 160 | 5.44 |
| Claude vs Publication | Red | Yellow | 194 | 6.59 |
| Claude vs Publication | Red | unknown | 16 | 0.54 |
| Claude vs Publication | Yellow | Green | 48 | 1.63 |
| Claude vs Publication | Yellow | Red | 831 | 28.25 |
| Claude vs Publication | Yellow | Yellow | 748 | 25.42 |
| Claude vs Publication | Yellow | unknown | 70 | 2.38 |
| Gemini vs Claude | Green | Green | 597 | 20.29 |
| Gemini vs Claude | Green | Red | 186 | 6.32 |
| Gemini vs Claude | Green | Yellow | 469 | 15.94 |
| Gemini vs Claude | Red | Green | 139 | 4.72 |
| Gemini vs Claude | Red | Red | 124 | 4.21 |
| Gemini vs Claude | Red | Yellow | 335 | 11.39 |
| Gemini vs Claude | Yellow | Green | 91 | 3.09 |
| Gemini vs Claude | Yellow | Red | 108 | 3.67 |
| Gemini vs Claude | Yellow | Yellow | 893 | 30.35 |
| Gemini vs Perplexity | Green | Green | 227 | 7.72 |
| Gemini vs Perplexity | Green | Red | 408 | 13.87 |
| Gemini vs Perplexity | Green | Yellow | 109 | 3.70 |
| Gemini vs Perplexity | Green | unknown | 508 | 17.27 |
| Gemini vs Perplexity | Red | Green | 59 | 2.01 |
| Gemini vs Perplexity | Red | Red | 193 | 6.56 |
| Gemini vs Perplexity | Red | Yellow | 89 | 3.03 |
| Gemini vs Perplexity | Red | unknown | 257 | 8.74 |
| Gemini vs Perplexity | Yellow | Green | 85 | 2.89 |
| Gemini vs Perplexity | Yellow | Red | 288 | 9.79 |
| Gemini vs Perplexity | Yellow | Yellow | 281 | 9.55 |
| Gemini vs Perplexity | Yellow | unknown | 438 | 14.89 |
| Gemini vs Publication | Green | Green | 214 | 7.27 |
| Gemini vs Publication | Green | Red | 374 | 12.71 |
| Gemini vs Publication | Green | Yellow | 613 | 20.84 |
| Gemini vs Publication | Green | unknown | 51 | 1.73 |
| Gemini vs Publication | Red | Green | 40 | 1.36 |
| Gemini vs Publication | Red | Red | 262 | 8.91 |
| Gemini vs Publication | Red | Yellow | 268 | 9.11 |
| Gemini vs Publication | Red | unknown | 28 | 0.95 |
| Gemini vs Publication | Yellow | Green | 17 | 0.58 |
| Gemini vs Publication | Yellow | Red | 555 | 18.86 |
| Gemini vs Publication | Yellow | Yellow | 479 | 16.28 |
| Gemini vs Publication | Yellow | unknown | 41 | 1.39 |
| OpenAI vs Claude | Green | Green | 717 | 24.37 |
| OpenAI vs Claude | Green | Red | 284 | 9.65 |
| OpenAI vs Claude | Green | Yellow | 1096 | 37.25 |
| OpenAI vs Claude | Red | Green | 86 | 2.92 |
| OpenAI vs Claude | Red | Red | 89 | 3.03 |
| OpenAI vs Claude | Red | Yellow | 199 | 6.76 |
| OpenAI vs Claude | Yellow | Green | 24 | 0.82 |
| OpenAI vs Claude | Yellow | Red | 45 | 1.53 |
| OpenAI vs Claude | Yellow | Yellow | 402 | 13.66 |
| OpenAI vs Gemini | Green | Green | 1054 | 35.83 |
| OpenAI vs Gemini | Green | Red | 388 | 13.19 |
| OpenAI vs Gemini | Green | Yellow | 655 | 22.26 |
| OpenAI vs Gemini | Red | Green | 139 | 4.72 |
| OpenAI vs Gemini | Red | Red | 115 | 3.91 |
| OpenAI vs Gemini | Red | Yellow | 120 | 4.08 |
| OpenAI vs Gemini | Yellow | Green | 59 | 2.01 |
| OpenAI vs Gemini | Yellow | Red | 95 | 3.23 |
| OpenAI vs Gemini | Yellow | Yellow | 317 | 10.77 |
| OpenAI vs Perplexity | Green | Green | 296 | 10.06 |
| OpenAI vs Perplexity | Green | Red | 647 | 21.99 |
| OpenAI vs Perplexity | Green | Yellow | 256 | 8.70 |
| OpenAI vs Perplexity | Green | unknown | 898 | 30.52 |
| OpenAI vs Perplexity | Red | Green | 50 | 1.70 |
| OpenAI vs Perplexity | Red | Red | 151 | 5.13 |
| OpenAI vs Perplexity | Red | Yellow | 67 | 2.28 |
| OpenAI vs Perplexity | Red | unknown | 106 | 3.60 |
| OpenAI vs Perplexity | Yellow | Green | 25 | 0.85 |
| OpenAI vs Perplexity | Yellow | Red | 91 | 3.09 |
| OpenAI vs Perplexity | Yellow | Yellow | 156 | 5.30 |
| OpenAI vs Perplexity | Yellow | unknown | 199 | 6.76 |
| OpenAI vs Publication | Green | Green | 230 | 7.82 |
| OpenAI vs Publication | Green | Red | 782 | 26.58 |
| OpenAI vs Publication | Green | Yellow | 1002 | 34.06 |
| OpenAI vs Publication | Green | unknown | 83 | 2.82 |
| OpenAI vs Publication | Red | Green | 35 | 1.19 |
| OpenAI vs Publication | Red | Red | 143 | 4.86 |
| OpenAI vs Publication | Red | Yellow | 173 | 5.88 |
| OpenAI vs Publication | Red | unknown | 23 | 0.78 |
| OpenAI vs Publication | Yellow | Green | 6 | 0.20 |
| OpenAI vs Publication | Yellow | Red | 266 | 9.04 |
| OpenAI vs Publication | Yellow | Yellow | 185 | 6.29 |
| OpenAI vs Publication | Yellow | unknown | 14 | 0.48 |
| Perplexity vs Claude | Green | Green | 162 | 5.51 |
| Perplexity vs Claude | Green | Red | 50 | 1.70 |
| Perplexity vs Claude | Green | Yellow | 159 | 5.40 |
| Perplexity vs Claude | Red | Green | 289 | 9.82 |
| Perplexity vs Claude | Red | Red | 146 | 4.96 |
| Perplexity vs Claude | Red | Yellow | 454 | 15.43 |
| Perplexity vs Claude | Yellow | Green | 36 | 1.22 |
| Perplexity vs Claude | Yellow | Red | 53 | 1.80 |
| Perplexity vs Claude | Yellow | Yellow | 390 | 13.26 |
| Perplexity vs Claude | unknown | Green | 340 | 11.56 |
| Perplexity vs Claude | unknown | Red | 169 | 5.74 |
| Perplexity vs Claude | unknown | Yellow | 694 | 23.59 |
| Perplexity vs Publication | Green | Green | 60 | 2.04 |
| Perplexity vs Publication | Green | Red | 130 | 4.42 |
| Perplexity vs Publication | Green | Yellow | 167 | 5.68 |
| Perplexity vs Publication | Green | unknown | 14 | 0.48 |
| Perplexity vs Publication | Red | Green | 72 | 2.45 |
| Perplexity vs Publication | Red | Red | 321 | 10.91 |
| Perplexity vs Publication | Red | Yellow | 446 | 15.16 |
| Perplexity vs Publication | Red | unknown | 50 | 1.70 |
| Perplexity vs Publication | Yellow | Green | 9 | 0.31 |
| Perplexity vs Publication | Yellow | Red | 266 | 9.04 |
| Perplexity vs Publication | Yellow | Yellow | 188 | 6.39 |
| Perplexity vs Publication | Yellow | unknown | 16 | 0.54 |
| Perplexity vs Publication | unknown | Green | 130 | 4.42 |
| Perplexity vs Publication | unknown | Red | 474 | 16.11 |
| Perplexity vs Publication | unknown | Yellow | 559 | 19.00 |
| Perplexity vs Publication | unknown | unknown | 40 | 1.36 |

**Table S2.** – Agreement Matrix across all Evidence Sources.

| badge_combo | combo_count |
| --- | --- |
| Atropos \| Open AI \| Gemini \| Perplexity \| Claude \| System |  |
| Green \| Green \| Green \| unknown \| Green \| Yellow | 68 |
| Green \| Green \| Yellow \| unknown \| Yellow \| Red | 58 |
| Green \| Green \| Green \| Red \| Green \| Yellow | 49 |
| Green \| Green \| Yellow \| unknown \| Yellow \| Yellow | 47 |
| Green \| Green \| Green \| unknown \| Yellow \| Yellow | 43 |
| Green \| Green \| Green \| unknown \| Green \| Green | 37 |
| Green \| Green \| Green \| unknown \| Yellow \| Red | 36 |
| Green \| Yellow \| Yellow \| unknown \| Yellow \| Red | 34 |
| Green \| Green \| Green \| Green \| Green \| Yellow | 30 |
| Red \| Green \| Yellow \| Yellow \| Yellow \| Red | 30 |
| Red \| Yellow \| Yellow \| Yellow \| Yellow \| Red | 29 |
| Red \| Green \| Yellow \| unknown \| Yellow \| Red | 28 |
| Green \| Green \| Green \| Red \| Yellow \| Yellow | 28 |
| Green \| Green \| Green \| Red \| Green \| Green | 28 |
| Green \| Green \| Green \| unknown \| Green \| Red | 28 |
| Green \| Green \| Yellow \| Red \| Yellow \| Yellow | 28 |
| Green \| Green \| Yellow \| Red \| Yellow \| Red | 27 |
| Red \| Green \| Yellow \| Red \| Yellow \| Red | 27 |
| Yellow \| Green \| Green \| Red \| Green \| Yellow | 27 |
| Green \| Green \| Red \| unknown \| Yellow \| Red | 27 |
| Green \| Green \| Red \| unknown \| Yellow \| Yellow | 25 |
| Green \| Yellow \| Yellow \| unknown \| Yellow \| Yellow | 24 |
| Red \| Green \| Yellow \| unknown \| Yellow \| Yellow | 24 |
| Green \| Green \| Green \| Green \| Green \| Green | 23 |
| Yellow \| Green \| Yellow \| unknown \| Yellow \| Red | 23 |
| Red \| Green \| Green \| unknown \| Green \| Yellow | 22 |
| Green \| Yellow \| Yellow \| Yellow \| Yellow \| Red | 21 |
| Yellow \| Green \| Yellow \| Yellow \| Yellow \| Red | 21 |
| Yellow \| Green \| Green \| unknown \| Green \| Yellow | 21 |
| Red \| Yellow \| Yellow \| unknown \| Yellow \| Red | 20 |
| Yellow \| Green \| Yellow \| unknown \| Yellow \| Yellow | 20 |
| Green \| Green \| Green \| Yellow \| Yellow \| Yellow | 19 |
| Yellow \| Green \| Yellow \| Red \| Yellow \| Yellow | 19 |
| Green \| Green \| Green \| Red \| Yellow \| Red | 18 |
| Yellow \| Green \| Green \| unknown \| Yellow \| Yellow | 18 |
| Green \| Green \| Green \| unknown \| Red \| Yellow | 18 |
| Green \| Green \| Green \| Red \| Green \| Red | 18 |
| Red \| Green \| Green \| unknown \| Yellow \| Red | 17 |
| Red \| Green \| Red \| unknown \| Yellow \| Yellow | 17 |
| Red \| Yellow \| Yellow \| Yellow \| Yellow \| Yellow | 17 |
| Green \| Green \| Red \| unknown \| Green \| Green | 16 |
| Red \| Green \| Green \| Red \| Yellow \| Yellow | 16 |
| Green \| Green \| Yellow \| Yellow \| Yellow \| Yellow | 16 |
| Green \| Green \| Red \| unknown \| Green \| Yellow | 16 |
| Red \| Green \| Green \| Red \| Green \| Yellow | 16 |
| Red \| Green \| Yellow \| Red \| Yellow \| Yellow | 16 |
| Yellow \| Green \| Green \| Red \| Green \| Red | 16 |
| Red \| Yellow \| Yellow \| unknown \| Yellow \| Yellow | 15 |
| Yellow \| Yellow \| Yellow \| unknown \| Yellow \| Red | 15 |
| Green \| Green \| Yellow \| Yellow \| Yellow \| Red | 15 |
| Yellow \| Green \| Green \| Red \| Yellow \| Yellow | 15 |
| Green \| Green \| Green \| Green \| Yellow \| Yellow | 15 |
| Green \| Green \| Red \| Red \| Green \| Yellow | 14 |
| Yellow \| Yellow \| Yellow \| Yellow \| Yellow \| Yellow | 14 |
| Yellow \| Green \| Yellow \| Yellow \| Yellow \| Yellow | 14 |
| Green \| Green \| Red \| Red \| Yellow \| Yellow | 13 |
| Green \| Green \| Green \| unknown \| Yellow \| Green | 13 |
| Green \| Green \| Yellow \| unknown \| Red \| Yellow | 13 |
| Red \| Green \| Green \| Green \| Green \| Red | 13 |
| Yellow \| Green \| Green \| unknown \| Yellow \| Red | 13 |
| Green \| Green \| Yellow \| Green \| Yellow \| Yellow | 13 |
| Red \| Green \| Green \| Green \| Green \| Yellow | 13 |
| Yellow \| Green \| Yellow \| Red \| Yellow \| Red | 13 |
| Yellow \| Green \| Green \| Red \| Yellow \| Red | 12 |
| Red \| Green \| Red \| unknown \| Yellow \| Red | 12 |
| Red \| Yellow \| Yellow \| Red \| Yellow \| Red | 12 |
| Green \| Green \| Yellow \| unknown \| Green \| Yellow | 12 |
| Green \| Green \| Red \| Red \| Yellow \| Red | 12 |
| Red \| Green \| Green \| Green \| Yellow \| Red | 12 |
| Green \| Green \| Green \| Green \| Yellow \| Red | 11 |
| Yellow \| Yellow \| Yellow \| Yellow \| Yellow \| Red | 11 |
| Red \| Green \| Yellow \| Yellow \| Yellow \| Yellow | 11 |
| Red \| Green \| Green \| unknown \| Green \| Green | 11 |
| Green \| Yellow \| Yellow \| Yellow \| Yellow \| Yellow | 11 |
| Red \| Green \| Green \| unknown \| Yellow \| Yellow | 11 |
| Green \| Green \| Yellow \| Green \| Yellow \| Red | 11 |
| Red \| Green \| Red \| Red \| Yellow \| Yellow | 10 |
| Green \| Yellow \| Red \| unknown \| Yellow \| Red | 10 |
| Red \| Green \| Green \| unknown \| Green \| Red | 10 |
| Red \| Yellow \| Yellow \| Red \| Yellow \| Yellow | 10 |
| Green \| Green \| Green \| Red \| Red \| Yellow | 10 |
| Green \| Green \| Green \| Green \| Green \| Red | 9 |
| Red \| Green \| Yellow \| Green \| Yellow \| Yellow | 9 |
| Green \| Green \| Yellow \| unknown \| Green \| Red | 9 |
| Green \| Yellow \| Yellow \| Red \| Yellow \| Yellow | 9 |
| Green \| Green \| Green \| Yellow \| Yellow \| Red | 9 |
| Red \| Green \| Green \| Red \| Red \| Yellow | 9 |
| Green \| Green \| Red \| unknown \| Red \| Yellow | 9 |
| Yellow \| Green \| Green \| Yellow \| Yellow \| Red | 9 |
| Red \| Green \| Yellow \| Red \| Green \| Yellow | 9 |
| Green \| Green \| Green \| Red \| Green \| unknown | 9 |
| Red \| Green \| Red \| Yellow \| Yellow \| Yellow | 8 |
| Yellow \| Red \| Yellow \| Yellow \| Yellow \| Red | 8 |
| Yellow \| Green \| Red \| Red \| Yellow \| Red | 8 |
| Green \| Green \| Red \| unknown \| Green \| Red | 8 |
| Red \| Green \| Green \| Yellow \| Yellow \| Yellow | 8 |
| Yellow \| Yellow \| Yellow \| Red \| Yellow \| Red | 8 |
| Green \| Red \| Red \| Red \| Green \| Yellow | 8 |
| Red \| Green \| Green \| Yellow \| Yellow \| Red | 8 |
| Red \| Green \| Green \| Green \| Green \| Green | 8 |
| Green \| Red \| Yellow \| Red \| Yellow \| Red | 8 |
| Green \| Green \| Green \| unknown \| Red \| Red | 8 |
| Yellow \| Green \| Green \| Green \| Green \| Yellow | 8 |
| Green \| Green \| Red \| unknown \| Red \| Red | 8 |
| Yellow \| Yellow \| Yellow \| unknown \| Yellow \| Yellow | 8 |
| Red \| Yellow \| Red \| Yellow \| Yellow \| Red | 7 |
| Yellow \| Green \| Green \| unknown \| Green \| Red | 7 |
| Green \| Red \| Green \| unknown \| Yellow \| Yellow | 7 |
| Red \| Green \| Red \| Red \| Red \| Yellow | 7 |
| Yellow \| Green \| Red \| Red \| Yellow \| Yellow | 7 |
| Red \| Red \| Red \| unknown \| Yellow \| Red | 7 |
| Yellow \| Green \| Green \| unknown \| Green \| Green | 7 |
| Green \| Green \| Red \| Red \| Red \| Red | 7 |
| Green \| Red \| Yellow \| Red \| Yellow \| Yellow | 7 |
| Yellow \| Yellow \| Yellow \| Red \| Yellow \| Yellow | 7 |
| Red \| Red \| Yellow \| Red \| Yellow \| Red | 7 |
| Green \| Green \| Yellow \| unknown \| Red \| Red | 7 |
| Yellow \| Green \| Green \| Red \| Green \| Green | 6 |
| Yellow \| Red \| Red \| Red \| Yellow \| Red | 6 |
| Yellow \| Green \| Yellow \| Green \| Yellow \| Yellow | 6 |
| Green \| Green \| Green \| Green \| Red \| Green | 6 |
| Green \| Red \| Green \| Red \| Green \| Yellow | 6 |
| Green \| Red \| Yellow \| unknown \| Yellow \| Yellow | 6 |
| Green \| Yellow \| Red \| unknown \| Yellow \| Yellow | 6 |
| Green \| Green \| Green \| unknown \| Red \| Green | 6 |
| Green \| Green \| Green \| unknown \| Yellow \| unknown | 6 |
| Red \| Yellow \| Red \| unknown \| Yellow \| Red | 6 |
| Red \| Green \| Green \| Red \| Yellow \| Red | 6 |
| Green \| Green \| Green \| Red \| Red \| Green | 6 |
| Green \| Green \| Green \| Red \| Red \| Red | 6 |
| Yellow \| Yellow \| Red \| Yellow \| Yellow \| Red | 6 |
| Yellow \| Green \| Green \| Red \| Red \| Red | 6 |
| Yellow \| Green \| Green \| Green \| Yellow \| Yellow | 6 |
| Red \| Green \| Red \| unknown \| Red \| Yellow | 6 |
| Red \| Red \| Yellow \| Yellow \| Yellow \| Red | 6 |
| Yellow \| Green \| Red \| unknown \| Green \| Yellow | 6 |
| Yellow \| Green \| Red \| Red \| Green \| Yellow | 6 |
| Yellow \| Green \| Green \| unknown \| Red \| Yellow | 6 |
| Red \| Green \| Red \| Red \| Yellow \| Red | 6 |
| Yellow \| Yellow \| Red \| unknown \| Yellow \| Red | 5 |
| Red \| Green \| Green \| unknown \| Red \| Red | 5 |
| Green \| Red \| Green \| unknown \| Green \| Yellow | 5 |
| Red \| Green \| Green \| unknown \| Red \| Yellow | 5 |
| Red \| Yellow \| Red \| Red \| Yellow \| Red | 5 |
| Yellow \| Green \| Red \| unknown \| Yellow \| Yellow | 5 |
| Green \| Red \| Red \| unknown \| Red \| Yellow | 5 |
| Yellow \| Green \| Green \| unknown \| Green \| unknown | 5 |
| Red \| Yellow \| Red \| unknown \| Yellow \| Yellow | 5 |
| Red \| Green \| Yellow \| Red \| Red \| Yellow | 5 |
| Red \| Green \| Yellow \| Yellow \| Red \| Red | 5 |
| Green \| Green \| Green \| Green \| Red \| Red | 5 |
| Green \| Red \| Red \| unknown \| Yellow \| Yellow | 5 |
| Red \| Green \| Green \| Red \| Red \| Red | 5 |
| Green \| Green \| Yellow \| Red \| Green \| Red | 5 |
| Green \| Green \| Red \| Yellow \| Yellow \| Red | 5 |
| Green \| Yellow \| Yellow \| Red \| Yellow \| Red | 5 |
| Red \| Green \| Red \| Red \| Green \| Yellow | 5 |
| Red \| Green \| Yellow \| Red \| Yellow \| unknown | 5 |
| Yellow \| Red \| Yellow \| Red \| Yellow \| Red | 5 |
| Yellow \| Green \| Red \| Yellow \| Yellow \| Red | 5 |
| Red \| Green \| Yellow \| Green \| Yellow \| Red | 5 |
| Yellow \| Red \| Green \| Red \| Green \| Yellow | 5 |
| Green \| Red \| Green \| Red \| Yellow \| Red | 5 |
| Yellow \| Green \| Red \| unknown \| Yellow \| Red | 5 |
| Green \| Red \| Green \| Green \| Green \| Yellow | 5 |
| Yellow \| Green \| Yellow \| unknown \| Red \| Yellow | 4 |
| Yellow \| Yellow \| Green \| Yellow \| Yellow \| Red | 4 |
| Red \| Red \| Green \| unknown \| Yellow \| Yellow | 4 |
| Green \| Green \| Red \| Yellow \| Yellow \| Yellow | 4 |
| Green \| Green \| Red \| Green \| Green \| Yellow | 4 |
| Green \| Green \| Green \| Yellow \| Red \| Yellow | 4 |
| Red \| Green \| Green \| Red \| Green \| Green | 4 |
| Green \| Red \| Yellow \| Yellow \| Yellow \| Yellow | 4 |
| Yellow \| Green \| Yellow \| Green \| Yellow \| Red | 4 |
| Green \| Green \| Yellow \| Red \| Yellow \| unknown | 4 |
| Green \| Red \| Red \| Yellow \| Yellow \| Red | 4 |
| Yellow \| Green \| Green \| unknown \| Red \| Red | 4 |
| Yellow \| Yellow \| Red \| Red \| Yellow \| Red | 4 |
| Red \| Green \| Green \| Red \| Green \| Red | 4 |
| Yellow \| Green \| Green \| unknown \| Red \| Green | 4 |
| Yellow \| Green \| Green \| Green \| Green \| Red | 4 |
| Green \| Green \| Green \| Red \| Yellow \| unknown | 4 |
| Red \| Green \| Red \| Red \| Red \| Red | 4 |
| Red \| Green \| Yellow \| unknown \| Red \| Red | 4 |
| Red \| Yellow \| Yellow \| Yellow \| Red \| Red | 4 |
| Green \| Yellow \| Yellow \| Green \| Yellow \| Red | 4 |
| Yellow \| Red \| Red \| Red \| Yellow \| Yellow | 4 |
| Green \| Green \| Green \| Green \| Red \| Yellow | 4 |
| Green \| Green \| Green \| Yellow \| Green \| Yellow | 4 |
| Yellow \| Green \| Green \| Yellow \| Yellow \| Yellow | 4 |
| Red \| Yellow \| Green \| unknown \| Green \| Red | 4 |
| Green \| Red \| Green \| Red \| Yellow \| Yellow | 4 |
| Green \| Red \| Yellow \| Yellow \| Yellow \| Red | 4 |
| Green \| Green \| Green \| Yellow \| Green \| Red | 4 |
| Red \| Green \| Yellow \| unknown \| Green \| Red | 4 |
| Red \| Red \| Yellow \| Red \| Yellow \| Yellow | 4 |
| Green \| Red \| Green \| Red \| Red \| Yellow | 4 |
| Red \| Red \| Red \| Red \| Yellow \| Red | 4 |
| Yellow \| Red \| Red \| unknown \| Yellow \| Red | 4 |
| Green \| Green \| Red \| Red \| Red \| Yellow | 4 |
| Red \| Green \| Red \| unknown \| Red \| Red | 4 |
| Red \| Green \| Red \| Green \| Yellow \| Yellow | 4 |
| Red \| Green \| Green \| Green \| Red \| Yellow | 4 |
| Green \| Green \| Red \| Red \| Green \| Red | 4 |
| Green \| Green \| Yellow \| Red \| Green \| Yellow | 4 |
| Green \| Yellow \| Yellow \| unknown \| Red \| Red | 4 |
| Green \| Red \| Yellow \| unknown \| Yellow \| Red | 4 |
| Green \| Red \| Green \| unknown \| Red \| Yellow | 4 |
| Yellow \| Green \| Green \| Green \| Green \| Green | 4 |
| Green \| Red \| Red \| Red \| Yellow \| Yellow | 4 |
| Red \| Red \| Yellow \| Yellow \| Red \| Red | 3 |
| Yellow \| Red \| Yellow \| unknown \| Yellow \| Yellow | 3 |
| Red \| Red \| Red \| Yellow \| Yellow \| Red | 3 |
| Red \| Green \| Red \| Green \| Red \| Red | 3 |
| Green \| Yellow \| Red \| Yellow \| Yellow \| Red | 3 |
| Green \| Yellow \| Green \| Yellow \| Yellow \| Red | 3 |
| Red \| Green \| Yellow \| unknown \| Red \| Yellow | 3 |
| Green \| Green \| Red \| Green \| Yellow \| Red | 3 |
| Green \| Green \| Red \| unknown \| Red \| Green | 3 |
| Yellow \| Green \| Green \| Red \| Yellow \| Green | 3 |
| Red \| Yellow \| Green \| unknown \| Yellow \| Yellow | 3 |
| Red \| Red \| Green \| Red \| Red \| Yellow | 3 |
| Green \| Green \| Green \| Red \| Yellow \| Green | 3 |
| Yellow \| Green \| Yellow \| Green \| Green \| Yellow | 3 |
| Yellow \| Green \| Red \| Red \| Red \| Yellow | 3 |
| Red \| Green \| Red \| Yellow \| Yellow \| Red | 3 |
| Yellow \| Green \| Yellow \| Red \| Red \| Red | 3 |
| Green \| Red \| Yellow \| unknown \| Red \| Yellow | 3 |
| Yellow \| Yellow \| Green \| unknown \| Yellow \| Yellow | 3 |
| Green \| Green \| Green \| Yellow \| Green \| Green | 3 |
| Green \| Red \| Red \| Red \| Red \| Red | 3 |
| Yellow \| Green \| Yellow \| unknown \| Green \| Yellow | 3 |
| Yellow \| Green \| Red \| Green \| Yellow \| Yellow | 3 |
| Green \| Green \| Yellow \| unknown \| Yellow \| unknown | 3 |
| Yellow \| Red \| Yellow \| Yellow \| Yellow \| Yellow | 3 |
| Green \| Yellow \| Green \| unknown \| Yellow \| Red | 3 |
| Yellow \| Green \| Yellow \| Green \| Green \| Red | 3 |
| Green \| Red \| Green \| Green \| Green \| Green | 3 |
| Yellow \| Green \| Green \| unknown \| Yellow \| Green | 3 |
| Red \| Yellow \| Green \| unknown \| Yellow \| Red | 3 |
| Yellow \| Green \| Green \| Green \| Red \| Yellow | 3 |
| Red \| Red \| Yellow \| Yellow \| Yellow \| Yellow | 3 |
| Yellow \| Red \| Red \| Yellow \| Yellow \| Yellow | 3 |
| Red \| Green \| Green \| unknown \| Yellow \| unknown | 3 |
| Red \| Yellow \| Red \| Yellow \| Red \| Red | 3 |
| Red \| Green \| Red \| unknown \| Green \| Yellow | 3 |
| Red \| Red \| Green \| Red \| Green \| Yellow | 3 |
| Green \| Green \| Green \| unknown \| Green \| unknown | 3 |
| Red \| Red \| Green \| Red \| Yellow \| Red | 3 |
| Green \| Yellow \| Yellow \| unknown \| Yellow \| unknown | 3 |
| Yellow \| Green \| Yellow \| Red \| Red \| Yellow | 3 |
| Red \| Red \| Green \| Red \| Green \| Red | 3 |
| Green \| Green \| Yellow \| Yellow \| Red \| Red | 3 |
| Red \| Green \| Yellow \| Red \| Green \| Red | 3 |
| Red \| Green \| Green \| Yellow \| Green \| Yellow | 3 |
| Yellow \| Red \| Red \| Red \| Red \| Yellow | 3 |
| Green \| Red \| Green \| Red \| Red \| Green | 3 |
| Green \| Red \| Green \| Green \| Yellow \| Red | 3 |
| Green \| Green \| Yellow \| Red \| Red \| Yellow | 3 |
| Yellow \| Red \| Green \| Red \| Red \| Yellow | 3 |
| Red \| Green \| Red \| Green \| Green \| Red | 3 |
| Green \| Red \| Green \| Red \| Red \| Red | 3 |
| Green \| Red \| Green \| Green \| Green \| Red | 3 |
| Red \| Green \| Red \| Green \| Red \| Yellow | 2 |
| Red \| Green \| Yellow \| Green \| Yellow \| unknown | 2 |
| Yellow \| Green \| Red \| Green \| Yellow \| Red | 2 |
| Green \| Red \| Green \| unknown \| Red \| Green | 2 |
| Yellow \| Yellow \| Red \| Green \| Yellow \| Yellow | 2 |
| Red \| Red \| Green \| Red \| Yellow \| Yellow | 2 |
| Green \| Red \| Red \| unknown \| Green \| Yellow | 2 |
| Yellow \| Yellow \| Yellow \| Yellow \| Yellow \| unknown | 2 |
| Green \| Green \| Yellow \| Green \| Yellow \| unknown | 2 |
| Green \| Green \| Yellow \| Red \| Red \| Red | 2 |
| Yellow \| Red \| Red \| unknown \| Green \| Yellow | 2 |
| Red \| Green \| Yellow \| Red \| Red \| unknown | 2 |
| Red \| Red \| Red \| Yellow \| Red \| Red | 2 |
| Green \| Green \| Red \| Yellow \| Green \| Yellow | 2 |
| Red \| Green \| Green \| unknown \| Red \| Green | 2 |
| Yellow \| Red \| Yellow \| Red \| Yellow \| Yellow | 2 |
| Green \| Yellow \| Yellow \| unknown \| Red \| Yellow | 2 |
| Yellow \| Green \| Green \| Yellow \| Green \| Yellow | 2 |
| Green \| Yellow \| Green \| unknown \| Green \| Green | 2 |
| Red \| Red \| Red \| Green \| Red \| Red | 2 |
| Red \| Green \| Green \| Green \| Red \| Red | 2 |
| Red \| Green \| Green \| Yellow \| Red \| Yellow | 2 |
| Green \| Yellow \| Yellow \| Green \| Yellow \| Yellow | 2 |
| Green \| Red \| Green \| unknown \| Green \| Green | 2 |
| Red \| Yellow \| Green \| Yellow \| Yellow \| Yellow | 2 |
| Yellow \| Green \| Yellow \| unknown \| Green \| Red | 2 |
| Green \| Red \| Green \| Yellow \| Yellow \| Yellow | 2 |
| Yellow \| Green \| Green \| Red \| Red \| Yellow | 2 |
| Green \| Red \| Yellow \| Red \| Red \| Yellow | 2 |
| Green \| Green \| Red \| Red \| Yellow \| unknown | 2 |
| Yellow \| Green \| Red \| Red \| Yellow \| unknown | 2 |
| Green \| Green \| Yellow \| Green \| Green \| Red | 2 |
| Green \| Yellow \| Red \| Red \| Yellow \| Yellow | 2 |
| Yellow \| Green \| Yellow \| Red \| Green \| Yellow | 2 |
| Yellow \| Red \| Green \| Yellow \| Red \| Yellow | 2 |
| Green \| Red \| Red \| Green \| Yellow \| Yellow | 2 |
| Green \| Red \| Green \| unknown \| Red \| Red | 2 |
| Green \| Red \| Yellow \| Green \| Yellow \| Yellow | 2 |
| Green \| Yellow \| Red \| Yellow \| Yellow \| Yellow | 2 |
| Green \| Yellow \| Green \| unknown \| Red \| Yellow | 2 |
| Red \| Green \| Yellow \| Yellow \| Yellow \| unknown | 2 |
| Yellow \| Red \| Red \| Green \| Yellow \| Red | 2 |
| Green \| Red \| Red \| Yellow \| Yellow \| Yellow | 2 |
| Red \| Yellow \| Red \| Yellow \| Yellow \| Yellow | 2 |
| Green \| Red \| Yellow \| unknown \| Green \| Red | 2 |
| Yellow \| Green \| Yellow \| Red \| Green \| Red | 2 |
| Green \| Green \| Red \| Red \| Green \| Green | 2 |
| Green \| Red \| Red \| Red \| Yellow \| Red | 2 |
| Green \| Red \| Yellow \| Red \| Green \| Yellow | 2 |
| Yellow \| Yellow \| Green \| Red \| Yellow \| Yellow | 2 |
| Green \| Green \| Red \| Green \| Red \| Red | 2 |
| Red \| Green \| Red \| unknown \| Green \| Green | 2 |
| Green \| Yellow \| Yellow \| Red \| Red \| Red | 2 |
| Yellow \| Red \| Red \| Yellow \| Yellow \| Red | 2 |
| Red \| Red \| Green \| Red \| Red \| Green | 2 |
| Green \| Red \| Red \| Yellow \| Red \| Red | 2 |
| Yellow \| Yellow \| Yellow \| unknown \| Red \| Yellow | 2 |
| Yellow \| Green \| Red \| Red \| Red \| unknown | 2 |
| Green \| Red \| Red \| Green \| Green \| Yellow | 2 |
| Yellow \| Green \| Red \| unknown \| Red \| Yellow | 2 |
| Yellow \| Green \| Red \| Green \| Green \| Red | 2 |
| Red \| Yellow \| Green \| Red \| Yellow \| Yellow | 2 |
| Green \| Red \| Green \| Green \| Red \| Green | 2 |
| Red \| Green \| Yellow \| Yellow \| Green \| Red | 2 |
| Green \| Red \| Green \| Green \| Yellow \| Yellow | 2 |
| Yellow \| Yellow \| Red \| unknown \| Yellow \| Yellow | 2 |
| Red \| Red \| Red \| Red \| Yellow \| Yellow | 2 |
| Green \| Red \| Green \| Red \| Green \| Red | 2 |
| Yellow \| Red \| Yellow \| Red \| Red \| Yellow | 2 |
| Yellow \| Green \| Green \| Yellow \| Red \| Yellow | 2 |
| Yellow \| Green \| Red \| Yellow \| Red \| Red | 2 |
| Red \| Green \| Red \| Yellow \| Red \| Red | 2 |
| Red \| Green \| Red \| unknown \| Green \| Red | 2 |
| Green \| Green \| Yellow \| Yellow \| Red \| Yellow | 2 |
| Green \| Red \| Yellow \| Yellow \| Green \| Yellow | 2 |
| Green \| Yellow \| Red \| unknown \| Red \| Yellow | 2 |
| Yellow \| Red \| Green \| Red \| Green \| Red | 2 |
| Green \| Red \| Green \| Red \| Green \| Green | 2 |
| Yellow \| Green \| Green \| Green \| Yellow \| Red | 2 |
| Red \| Green \| Red \| Red \| Green \| Red | 2 |
| Green \| Green \| Yellow \| Yellow \| Green \| Yellow | 2 |
| Red \| Yellow \| Red \| unknown \| Red \| Red | 2 |
| Red \| Green \| Green \| Green \| Green \| unknown | 2 |
| Red \| Green \| Red \| Red \| Green \| Green | 2 |
| Red \| Yellow \| Green \| Red \| Green \| Yellow | 2 |
| Red \| Yellow \| Green \| Green \| Yellow \| Yellow | 2 |
| Green \| Yellow \| Yellow \| unknown \| Yellow \| Green | 2 |
| Green \| Yellow \| Green \| Red \| Yellow \| Red | 2 |
| Green \| Green \| Green \| Green \| Yellow \| Green | 2 |
| Green \| Green \| Red \| Red \| Green \| unknown | 2 |
| Green \| Red \| Yellow \| unknown \| Yellow \| Green | 2 |
| Yellow \| Red \| Green \| unknown \| Yellow \| Red | 2 |
| Green \| Yellow \| Green \| Yellow \| Green \| Red | 1 |
| Yellow \| Green \| Green \| Green \| Red \| Green | 1 |
| Yellow \| Yellow \| Green \| Green \| Red \| Yellow | 1 |
| Red \| Yellow \| Red \| Green \| Green \| Red | 1 |
| Yellow \| Green \| Red \| Yellow \| Green \| Yellow | 1 |
| Yellow \| Red \| Green \| unknown \| Yellow \| Yellow | 1 |
| Red \| Green \| Red \| unknown \| Red \| Green | 1 |
| Green \| Red \| Green \| unknown \| Yellow \| Red | 1 |
| Red \| Green \| Green \| Green \| Yellow \| Yellow | 1 |
| Green \| Red \| Red \| Green \| Red \| unknown | 1 |
| Yellow \| Green \| Red \| Red \| Red \| Red | 1 |
| Green \| Red \| Red \| Green \| Red \| Green | 1 |
| Yellow \| Red \| Green \| Red \| Red \| Red | 1 |
| Yellow \| Red \| Green \| Red \| Yellow \| Green | 1 |
| Red \| Red \| Yellow \| unknown \| Yellow \| Red | 1 |
| Red \| Green \| Red \| Green \| Yellow \| unknown | 1 |
| Yellow \| Green \| Green \| Yellow \| Green \| Red | 1 |
| Red \| Green \| Red \| Yellow \| Yellow \| unknown | 1 |
| Red \| Red \| Red \| unknown \| Green \| Yellow | 1 |
| Green \| Yellow \| Red \| Red \| Green \| unknown | 1 |
| Green \| Red \| Yellow \| Yellow \| Red \| Yellow | 1 |
| Green \| Red \| Green \| unknown \| Yellow \| Green | 1 |
| Yellow \| Yellow \| Yellow \| unknown \| Red \| Red | 1 |
| Green \| Green \| Red \| Red \| Red \| Green | 1 |
| Green \| Yellow \| Red \| Red \| Red \| unknown | 1 |
| Yellow \| Red \| Red \| Red \| Green \| Red | 1 |
| Green \| Yellow \| Green \| unknown \| Yellow \| Yellow | 1 |
| Green \| Green \| Red \| Yellow \| Green \| Red | 1 |
| Red \| Green \| Green \| Red \| Green \| unknown | 1 |
| Green \| Green \| Green \| Yellow \| Red \| Red | 1 |
| Red \| Yellow \| Yellow \| Green \| Yellow \| Yellow | 1 |
| Red \| Red \| Green \| unknown \| Green \| Green | 1 |
| Red \| Yellow \| Red \| unknown \| Yellow \| unknown | 1 |
| Green \| Red \| Yellow \| unknown \| Green \| Yellow | 1 |
| Yellow \| Green \| Red \| Green \| Yellow \| Green | 1 |
| Red \| Yellow \| Yellow \| Yellow \| Red \| Yellow | 1 |
| Yellow \| Red \| Green \| Red \| Yellow \| unknown | 1 |
| Yellow \| Green \| Yellow \| Yellow \| Red \| Green | 1 |
| Red \| Yellow \| Red \| Red \| Yellow \| unknown | 1 |
| Green \| Yellow \| Green \| Red \| Red \| Red | 1 |
| Green \| Yellow \| Green \| Yellow \| Yellow \| Green | 1 |
| Red \| Green \| Green \| Yellow \| Yellow \| unknown | 1 |
| Yellow \| Yellow \| Green \| Red \| Green \| Yellow | 1 |
| Yellow \| Green \| Yellow \| unknown \| Red \| Red | 1 |
| Green \| Green \| Yellow \| Yellow \| Yellow \| unknown | 1 |
| Yellow \| Yellow \| Green \| unknown \| Yellow \| Red | 1 |
| Red \| Yellow \| Yellow \| unknown \| Red \| Red | 1 |
| Red \| Yellow \| Red \| unknown \| Green \| unknown | 1 |
| Green \| Red \| Red \| Green \| Yellow \| Red | 1 |
| Green \| Green \| Yellow \| unknown \| Green \| unknown | 1 |
| Red \| Red \| Red \| Red \| Green \| unknown | 1 |
| Yellow \| Green \| Red \| unknown \| Green \| Green | 1 |
| Red \| Green \| Yellow \| unknown \| Yellow \| Green | 1 |
| Yellow \| Red \| Yellow \| Green \| Green \| Yellow | 1 |
| Red \| Red \| Yellow \| Green \| Yellow \| Yellow | 1 |
| Red \| Yellow \| Red \| Red \| Yellow \| Yellow | 1 |
| Yellow \| Red \| Red \| Yellow \| Yellow \| unknown | 1 |
| Yellow \| Red \| Red \| unknown \| Red \| unknown | 1 |
| Yellow \| Green \| Red \| Yellow \| Green \| Red | 1 |
| Yellow \| Yellow \| Red \| unknown \| Red \| Red | 1 |
| Red \| Yellow \| Yellow \| unknown \| Yellow \| unknown | 1 |
| Red \| Red \| Green \| Red \| Red \| unknown | 1 |
| Green \| Green \| Yellow \| Red \| Green \| Green | 1 |
| Red \| Green \| Green \| Red \| Red \| Green | 1 |
| Red \| Yellow \| Red \| Red \| Red \| Yellow | 1 |
| Yellow \| Yellow \| Yellow \| Yellow \| Red \| Red | 1 |
| Green \| Yellow \| Green \| Red \| Yellow \| Yellow | 1 |
| Red \| Green \| Yellow \| Red \| Red \| Red | 1 |
| Red \| Red \| Red \| Yellow \| Red \| Green | 1 |
| Green \| Green \| Red \| Yellow \| Red \| Red | 1 |
| Red \| Green \| Red \| Yellow \| Green \| Red | 1 |
| Yellow \| Red \| Green \| unknown \| Red \| Red | 1 |
| Green \| Green \| Yellow \| Green \| Yellow \| Green | 1 |
| Red \| Yellow \| Yellow \| Yellow \| Green \| Red | 1 |
| Yellow \| Red \| Green \| Yellow \| Yellow \| Yellow | 1 |
| Green \| Green \| Red \| Yellow \| Red \| Yellow | 1 |
| Green \| Green \| Yellow \| unknown \| Red \| unknown | 1 |
| Red \| Yellow \| Red \| Red \| Red \| Red | 1 |
| Yellow \| Red \| Yellow \| Yellow \| Yellow \| unknown | 1 |
| Yellow \| Red \| Green \| unknown \| Yellow \| Green | 1 |
| Red \| Green \| Yellow \| Green \| Red \| Red | 1 |
| Yellow \| Red \| Yellow \| Green \| Yellow \| Red | 1 |
| Red \| Green \| Green \| unknown \| Yellow \| Green | 1 |
| Green \| Red \| Yellow \| Green \| Red \| Yellow | 1 |
| Red \| Green \| Green \| Red \| Yellow \| unknown | 1 |
| Green \| Yellow \| Red \| Red \| Yellow \| Red | 1 |
| Yellow \| Yellow \| Green \| Green \| Green \| Yellow | 1 |
| Yellow \| Red \| Yellow \| Red \| Red \| unknown | 1 |
| Green \| Yellow \| Red \| Green \| Green \| Yellow | 1 |
| Green \| Yellow \| Yellow \| Yellow \| Yellow \| unknown | 1 |
| Green \| Green \| Red \| Green \| Yellow \| Yellow | 1 |
| Yellow \| Green \| Yellow \| unknown \| Yellow \| Green | 1 |
| Green \| Red \| Yellow \| Green \| Yellow \| Green | 1 |
| Red \| Green \| Yellow \| unknown \| Green \| Yellow | 1 |
| Green \| Yellow \| Yellow \| Green \| Green \| Yellow | 1 |
| Yellow \| Red \| Green \| Green \| Green \| Green | 1 |
| Red \| Green \| Green \| Green \| Yellow \| Green | 1 |
| Yellow \| Green \| Red \| Yellow \| Yellow \| Yellow | 1 |
| Yellow \| Green \| Red \| Red \| Green \| Green | 1 |
| Red \| Red \| Green \| unknown \| Yellow \| Red | 1 |
| Red \| Green \| Red \| Green \| Green \| Green | 1 |
| Green \| Red \| Red \| Yellow \| Yellow \| unknown | 1 |
| Yellow \| Green \| Red \| Red \| Red \| Green | 1 |
| Yellow \| Yellow \| Green \| Red \| Yellow \| unknown | 1 |
| Red \| Red \| Green \| Green \| Green \| Yellow | 1 |
| Green \| Green \| Red \| Green \| Yellow \| unknown | 1 |
| Yellow \| Green \| Green \| Red \| Yellow \| unknown | 1 |
| Red \| Red \| Red \| Green \| Green \| Red | 1 |
| Green \| Green \| Red \| Green \| Yellow \| Green | 1 |
| Yellow \| Yellow \| Yellow \| Red \| Red \| Red | 1 |
| Yellow \| Yellow \| Red \| Red \| Red \| Yellow | 1 |
| Red \| Red \| Yellow \| Green \| Yellow \| Red | 1 |
| Green \| Yellow \| Yellow \| unknown \| Green \| Yellow | 1 |
| Red \| Yellow \| Red \| Green \| Yellow \| Red | 1 |
| Yellow \| Red \| Green \| unknown \| Green \| Yellow | 1 |
| Red \| Red \| Yellow \| Red \| Red \| Yellow | 1 |
| Red \| Red \| Yellow \| unknown \| Yellow \| Yellow | 1 |
| Yellow \| Green \| Green \| Green \| Yellow \| Green | 1 |
| Yellow \| Red \| Green \| unknown \| Green \| Red | 1 |
| Green \| Red \| Green \| unknown \| Yellow \| unknown | 1 |
| Yellow \| Red \| Green \| Green \| Green \| unknown | 1 |
| Green \| Red \| Green \| Red \| Green \| unknown | 1 |
| Red \| Green \| Red \| Green \| Yellow \| Red | 1 |
| Yellow \| Yellow \| Green \| Green \| Yellow \| Red | 1 |
| Red \| Red \| Green \| Green \| Red \| Yellow | 1 |
| Green \| Yellow \| Red \| Green \| Yellow \| Red | 1 |
| Yellow \| Yellow \| Yellow \| Red \| Red \| Yellow | 1 |
| Yellow \| Red \| Green \| Red \| Red \| Green | 1 |
| Red \| Red \| Green \| Green \| Green \| Green | 1 |
| Green \| Yellow \| Yellow \| unknown \| Green \| Red | 1 |
| Yellow \| Yellow \| Green \| Green \| Yellow \| Yellow | 1 |
| Red \| Red \| Green \| Red \| Red \| Red | 1 |
| Green \| Green \| Yellow \| unknown \| Yellow \| Green | 1 |
| Green \| Yellow \| Green \| Green \| Green \| Yellow | 1 |
| Yellow \| Green \| Yellow \| Red \| Yellow \| unknown | 1 |
| Yellow \| Red \| Yellow \| unknown \| Red \| Red | 1 |
| Green \| Green \| Green \| Yellow \| Yellow \| Green | 1 |
| Red \| Green \| Green \| Yellow \| Yellow \| Green | 1 |
| Green \| Red \| Yellow \| Yellow \| Yellow \| unknown | 1 |
| Red \| Green \| Yellow \| Green \| Green \| Yellow | 1 |
| Green \| Red \| Red \| unknown \| Red \| Green | 1 |
| Green \| Red \| Red \| Red \| Red \| Yellow | 1 |
| Yellow \| Green \| Red \| Yellow \| Yellow \| unknown | 1 |
| Red \| Green \| Yellow \| Green \| Red \| Yellow | 1 |
| Green \| Yellow \| Green \| Red \| Red \| Yellow | 1 |
| Green \| Yellow \| Green \| Yellow \| Yellow \| Yellow | 1 |
| Yellow \| Yellow \| Yellow \| Green \| Yellow \| Red | 1 |
| Green \| Red \| Green \| Yellow \| Yellow \| Red | 1 |
| Red \| Yellow \| Yellow \| Yellow \| Green \| Yellow | 1 |
| Yellow \| Red \| Yellow \| Red \| Green \| Yellow | 1 |
| Yellow \| Green \| Red \| unknown \| Red \| Red | 1 |
| Green \| Red \| Yellow \| unknown \| Yellow \| unknown | 1 |
| Yellow \| Red \| Red \| Yellow \| Red \| Red | 1 |
| Yellow \| Red \| Green \| Red \| Red \| unknown | 1 |
| Green \| Red \| Yellow \| Red \| Red \| Red | 1 |
| Yellow \| Red \| Red \| unknown \| Red \| Green | 1 |
| Yellow \| Red \| Red \| unknown \| Red \| Red | 1 |
| Green \| Green \| Yellow \| Red \| Yellow \| Green | 1 |
| Green \| Red \| Red \| Green \| Green \| Red | 1 |
| Green \| Yellow \| Green \| unknown \| Green \| Red | 1 |
| Green \| Red \| Green \| Yellow \| Green \| Yellow | 1 |
| Green \| Green \| Green \| Red \| Red \| unknown | 1 |
| Red \| Green \| Green \| Green \| Yellow \| unknown | 1 |
| Yellow \| Yellow \| Red \| Red \| Yellow \| Yellow | 1 |
| Green \| Green \| Red \| Green \| Green \| Red | 1 |
| Red \| Red \| Red \| Red \| Green \| Red | 1 |
| Green \| Yellow \| Green \| Green \| Red \| Red | 1 |
| Green \| Red \| Yellow \| unknown \| Green \| Green | 1 |
| Red \| Red \| Yellow \| Red \| Red \| unknown | 1 |
| Yellow \| Red \| Green \| unknown \| Green \| unknown | 1 |
| Green \| Red \| Green \| unknown \| Green \| unknown | 1 |
| Red \| Red \| Green \| Green \| Red \| Red | 1 |
| Red \| Red \| Red \| unknown \| Yellow \| Yellow | 1 |
| Green \| Green \| Green \| Green \| Yellow \| unknown | 1 |
| Yellow \| Green \| Green \| Yellow \| Red \| Red | 1 |
| Green \| Red \| Red \| Red \| Green \| Green | 1 |
| Yellow \| Red \| Red \| Red \| Red \| unknown | 1 |
| Green \| Red \| Red \| unknown \| Yellow \| Red | 1 |
| Green \| Yellow \| Red \| unknown \| Red \| Red | 1 |
| Green \| Green \| Red \| Green \| Red \| Yellow | 1 |
| Yellow \| Yellow \| Yellow \| Yellow \| Red \| Yellow | 1 |
| Red \| Red \| Green \| unknown \| Green \| Yellow | 1 |
| Green \| Green \| Yellow \| unknown \| Green \| Green | 1 |
| Green \| Yellow \| Yellow \| Yellow \| Red \| Red | 1 |
| Yellow \| Yellow \| Green \| Yellow \| Green \| Yellow | 1 |
| Green \| Red \| Red \| Green \| Red \| Yellow | 1 |
| Red \| Yellow \| Yellow \| Red \| Yellow \| Green | 1 |
| Green \| Green \| Yellow \| Green \| Red \| Yellow | 1 |
| Red \| Yellow \| Yellow \| Green \| Red \| Red | 1 |
| Yellow \| Red \| Yellow \| unknown \| Yellow \| unknown | 1 |
| Red \| Red \| Red \| Red \| Red \| Yellow | 1 |
| Red \| Red \| Yellow \| unknown \| Red \| Yellow | 1 |
| Green \| Yellow \| Red \| Yellow \| Yellow \| unknown | 1 |
| Green \| Red \| Green \| Green \| Red \| Red | 1 |
| Red \| Red \| Green \| Yellow \| Yellow \| Red | 1 |
| Yellow \| Green \| Yellow \| Yellow \| Green \| Red | 1 |
| Yellow \| Green \| Yellow \| Yellow \| Yellow \| unknown | 1 |
| Red \| Yellow \| Yellow \| Red \| Red \| Red | 1 |
| Yellow \| Green \| Yellow \| Yellow \| Green \| Yellow | 1 |
| Green \| Green \| Green \| Green \| Green \| unknown | 1 |
| Green \| Green \| Yellow \| Yellow \| Yellow \| Green | 1 |
| Yellow \| Red \| Green \| unknown \| Red \| Green | 1 |
| Green \| Green \| Red \| unknown \| Yellow \| Green | 1 |
| Green \| Green \| Red \| unknown \| Green \| unknown | 1 |
| Yellow \| Red \| Yellow \| unknown \| Yellow \| Red | 1 |
| Yellow \| Red \| Yellow \| Green \| Yellow \| Yellow | 1 |
| Yellow \| Green \| Yellow \| unknown \| Green \| Green | 1 |
| Green \| Red \| Red \| Yellow \| Red \| unknown | 1 |
| Yellow \| Green \| Yellow \| Red \| Green \| unknown | 1 |
| Yellow \| Green \| Red \| Red \| Green \| Red | 1 |
| Red \| Yellow \| Red \| Red \| Green \| Red | 1 |
| Green \| Green \| Red \| Green \| Green \| unknown | 1 |
| Red \| Yellow \| Green \| Yellow \| Yellow \| Red | 1 |
| Red \| Green \| Green \| unknown \| Red \| unknown | 1 |
| Green \| Red \| Yellow \| Green \| Yellow \| Red | 1 |
| Red \| Green \| Yellow \| unknown \| Yellow \| unknown | 1 |
| Green \| Yellow \| Green \| Green \| Yellow \| Yellow | 1 |
| Yellow \| Green \| Green \| unknown \| Yellow \| unknown | 1 |
| Green \| Red \| Red \| unknown \| Yellow \| unknown | 1 |
| Yellow \| Yellow \| Red \| Yellow \| Red \| Red | 1 |
| Yellow \| Red \| Green \| Red \| Green \| Green | 1 |
| Red \| Red \| Yellow \| unknown \| Yellow \| unknown | 1 |
| Red \| Red \| Red \| Green \| Green \| Green | 1 |
| Red \| Red \| Red \| unknown \| Red \| Yellow | 1 |
| Green \| Yellow \| Red \| Yellow \| Red \| Red | 1 |

